# ICON (Ivermectin in COvid Nineteen) study: Use of Ivermectin is Associated with Lower Mortality in Hospitalized Patients with COVID19

**DOI:** 10.1101/2020.06.06.20124461

**Authors:** Juliana Cepelowicz Rajter, Michael S. Sherman, Naaz Fatteh, Fabio Vogel, Jamie Sacks, Jean-Jacques Rajter

## Abstract

**Importance:** No therapy to date has been shown to improve survival for patients infected with SARS-CoV-2. Ivermectin has been shown to inhibit the replication of SARS-CoV-2 in vitro but clinical response has not been previously evaluated.

**Objective:** To determine whether Ivermectin is associated with lower mortality rate in patients hospitalized with COVID-19.

**Design and Setting:** Retrospective cohort study of consecutive patients hospitalized at four Broward Health hospitals in South Florida with confirmed SARS-CoV-2. Enrollment dates were March 15, 2020 through May 11, 2020. Follow up data for all outcomes was May 19, 2020.

**Participants:** 280 patients with confirmed SARS-CoV-2 infection (mean age 59.6 years [standard deviation 17.9], 45.4% female), of whom 173 were treated with ivermectin and 107 were usual care were reviewed. 27 identified patients were not reviewed due to multiple admissions, lack of confirmed COVID results during hospitalization, age less than 18, pregnancy, or incarceration.

**Exposure:** Patients were categorized into two treatment groups based on whether they received at least one dose of ivermectin at any time during the hospitalization. Treatment decisions were at the discretion of the treating physicians. Severe pulmonary involvement at study entry was characterized as need for either FiO2 ≥50%, or noninvasive or invasive mechanical ventilation.

**Main Outcomes and Measures:** The primary outcome was all-cause in-hospital mortality. Secondary outcomes included subgroup mortality in patients with severe pulmonary involvement and extubation rates for patients requiring invasive ventilation.

**Results:** Univariate analysis showed lower mortality in the ivermectin group (15.0% versus 25.2%, OR 0.52, 95% CI 0.29-0.96, P=.03). Mortality was also lower among 75 patients with severe pulmonary disease treated with ivermectin (38.8% vs 80.7%, OR 0.15, CI 0.05-0.47, P=.001), but there was no significant difference in successful extubation rates (36.1% vs 15.4%, OR 3.11 (0.88-11.00), p=.07). After adjustment for between-group differences and mortality risks, the mortality difference remained significant for the entire cohort (OR 0.27, CI 0.09-0.85, p=.03; HR 0.37, CI 0.19-0.71, p=.03).

**Conclusions and Relevance:** Ivermectin was associated with lower mortality during treatment of COVID-19, especially in patients who required higher inspired oxygen or ventilatory support. These findings should be further evaluated with randomized controlled trials.

## Introduction

As of May 24th, 2020, the CDC recorded nearly 100,000 people having died of Covid-19 with at least 1,639,099 confirmed infections having been reported in the US and its territories. Covid-19 presents an unprecedented challenge to identify effective therapy for prevention and treatment. Currently, there is no evidence from randomized controlled trials of any potential therapy improving survival outcomes in patients with confirmed disease.

In the late 1970s Ivermectin was developed as a new class of drug to treat parasitic infections. Initially used in veterinary Medicine, it was soon found to be safe and effective in humans. It has successfully been used to treat onchocerciasis and lymphatic filariasis in millions of people worldwide as part of a global drug donation program. About 3.7 billion doses of Ivermectin have been distributed in mass drug administration campaigns globally over the past 30 years. Presently, Ivermectin is approved for use in humans in several countries to treat onchocerciasis, lymphatic filariasis, strongyloidiasis and scabies.^1^

Ivermectin has previously been studied as a therapeutic option for viral infections with in vitro data showing some activity against a broad range of viruses, including HIV, Dengue, Influenza and Zika virus.^2,3^ In a recent study, Wagstaff et al, demonstrated that Ivermectin was a potent in-vitro inhibitor of SARS-CoV-2, showing a 99.8% reduction in viral RNA after 48 hours.^4^ However, in-vivo efficacy of ivermectin in SARS-CoV-2 infection in humans has not previously been reported.

## Methods

### Patients

Sequentially consecutive hospitalized patients at four Broward Health associated hospitals in South Florida with laboratory-confirmed infection with SARS-CoV-2 during their admission were reviewed in this study. The list of confirmed cases was provided by the epidemiology department. Enrollment dates ranged from March 15, 2020 through May 11, 2020. Confirmatory testing was performed by nasopharyngeal swab using an FDA Emergency Use Authorized COVID-19 molecular assay for the detection of SARS-CoV-2 RNA. Patients younger than 18 years old, pregnant, or incarcerated were excluded from the data collection based on IRB requirements. Patients who had at least 2 separate admissions placing them in both groups were also excluded.

### Trial procedures

Records were abstracted by four of the authors and all data was subsequently reviewed and confirmed by the lead author. Baseline data was collected at the time of ivermectin administration for the ivermectin group; for the usual care group baseline was either at the time of administration of hydroxychloroquine or, if not used, at the time of admission. Information collected included COVID-19 testing results, patient demographics, pre-existing comorbid conditions, initial vital signs, chest imaging studies, laboratory results, and the use of hydroxychloroquine with and without azithromycin in order to describe the cohort and to identify potential cofounders between groups. Severity of pulmonary involvement was assessed at the time of initiation of therapy (“onset”) and categorized as severe or non-severe. Patients were considered to have severe pulmonary involvement if they required an FiO2 of 50% or greater, high-flow nasal oxygen, noninvasive ventilation, or intubation and mechanical ventilation. The non-severe pulmonary criteria encompassed patients who required no supplemental oxygen, or “low FIO2” (ie: Venturi mask 40% or less, or any amount of low flow nasal cannula), independent of radiographic or laboratory findings.

Patients were categorized into two treatment groups based on whether they received ivermectin at any time during the hospitalization. Patients in the Ivermectin group received at least one oral dose of ivermectin at 200 micrograms/kilogram in addition to usual clinical care. The decision to prescribe ivermectin, hydroxychloroquine, azithromycin or other medications was at the discretion of the treating physicians, however hospital guidelines were established for the use of these agents as well as for cardiac and QT monitoring for patients receiving hydroxychloroquine. Oxygen and ventilatory support were applied per the customary care.

### Outcomes

The primary outcome was all-cause in-hospital mortality. Patient was considered a “survivor” if they left the hospital alive, or if their status in the hospital changed from active care to awaiting transfer to a skilled facility. The latter outcome was necessitated by the requirement that two consecutive negative nasopharyngeal swab specimens for SARS-CoV-2, collected equal to or greater than 24 hours apart, were necessary for a patient to be accepted to a skilled nursing facility.

Secondary outcomes included subgroup mortality of patients with severe pulmonary involvement, extubation rates for patients requiring mechanical ventilation, and length of hospital stay.

### Statistical analysis

Univariate analysis of the primary mortality outcome, and comparisons between treatment groups were determined by Student’s t test or Mann-Whitney U test for continuous variables as appropriate, and by Pearson Chi Square test for categorical variables. The method of Hodges-Lehman was used to estimate median differences with 95% confidence intervals.

To adjust for confounders and between-group differences, a multivariate analysis was performed using stepwise binary logistic regression. Patient variables included in the analysis were age, sex, comorbidities of diabetes, chronic lung disease, cardiovascular disease, and hypertension, smoking status, severity of pulmonary involvement, BMI, peripheral white blood count, absolute lymphocyte count, and use of hydroxychloroquine and azithromycin based on bivariate associations within our data, a priori plausibility, and documented associations with mortality from previous studies.

Adjusted odds ratio with 95% confidence intervals were computed to show level of certainty. Hazard ratios for the primary outcome of mortality with 95% confidence intervals were calculated by means of the Cox regression with the same covariates. Analyses were based on nonmissing data and missing data were not imputed. Missingness of 1% was found for peripheral white blood cell count, 5% for smoking status, and 7% for absolute lymphocyte count. Secondary analyses were thus also performed on the entire cohort. All tests were 2-sided and a p value <.05 was considered statistically significant. All statistical analyses were conducted using IBM SPSS v 24.0 software.

### Oversight

The protocol was approved by the institutional review board for the Broward Health hospital system. The authors assume responsibility for the accuracy and completeness of the data and analyses, as well as for the fidelity of the trial.

## Results

### Characteristics of the patients

307 patients were admitted for COVID-19 during the time period studied. 4 patients were not reviewed due to multiple admissions, 11 had no confirmed COVID testing at the time of the study, and 12 were excluded due to age younger than 18 years old, pregnancy, or incarceration. The remaining cohort of 280 patients was comprised of 173 treated with ivermectin and 107 in the usual care group. Follow up data for all outcomes were available through May 19th, 2020. No patients were lost to follow-up for the primary outcome. At the time of analysis, all patients in the cohort had met the endpoint of death, discharge alive, or awaiting transfer to a skilled facility.

Baseline characteristics and between group comparisons are shown in Table 1. Characteristics were similar between groups, however hypertension was more prevalent in the Ivermectin group, whereas the use of hydroxychloroquine and hydroxychloroquine plus azithromycin were higher in the usual care group. No other significant between-group differences were found among baseline characteristics or comorbidities, including age, race, cardiac comorbidities, or smoking status.

**Table 1:** Patient Characteristics by Treatment Group

| Demographic characteristics | Total (n=280) | Usual Care (n=107) | Ivermectin (n=173) | P value |
| --- | --- | --- | --- | --- |
| Age, years<sup>a</sup> | 59.6 (17.9) | 58.6 (18.5) | 60.2 (17.6) | .45 |
| Female sex | 127 (45.4) | 43 (41.2) | 84 (48.6) | .17 |
| Race ethnicity |  |  |  | .36 |
| Black | 153 (54.6) | 55 (51.4) | 98 (56.6) |  |
| White | 76 (27.1) | 35 (32.7) | 41 (23.7) |  |
| Hispanic | 33 (11.7) | 12 (11.2) | 21 (12.1) |  |
| Other or not identified<sup>c</sup> | 13 (4.6) | 5 (4.7) | 13 (7.5) |  |
| Smoking |  |  |  | .40 |
| Current | 16/255 (6.3) | 7/99 (7.1) | 9/156 (5.7) |  |
| Former | 30/255 (11.7) | 15/99 (15.2) | 15/151 (9.9) |  |
| Never | 209/255 (81.9) | 77/99 (77.8) | 132/151 (87.4) |  |
| Number of comorbidities<sup>a</sup> | 1.66 (1.34) | 1.60 (1.46) | 1.70 (1.27) | .57 |
| Diabetes | 90 (32.1) | 31 (29.0) | 59 (34.1) | .37 |
| Cardiac | 43 (15.4) | 18 (16.8) | 25 (14.5) | .59 |
| Pulmonary | 28 (10.0) | 14 (13.1) | 14 (8.9) | .18 |
| Obesity | 114 (40.7) | 42 (39.3) | 72 (41.6) | .70 |
| Renal | 24 (8.6) | 10 (9.4) | 14 (8.1) | .72 |
| Cancer | 17 (6.1) | 8 (7.5) | 9 (5.2) | .44 |
| Hypertension | 50 (17.9) | 13 (12.2) | 37 (21.4) | .05 |
| Neurologic | 28 (10.0) | 8 (7.5) | 20 (11.6) | .27 |
| HIV | 9 (3.2) | 1 (1) | 8 (4.6) | .09 |
| Thyroid | 23 (8.2) | 7 (6.6) | 16 (9.3) | .42 |
| BMI<sup>a</sup> | 30.0 (7.8) | 29.8 (7.2) | 30.1 (8.2) | .81 |
| Severity |  |  |  | .46 |
| Severe | 75 (26.8) | 26 (24.3) | 49 (28.3) | .12 |
| Non-severe | 205 (73.2) | 81 (75.7) | 124 (71.7) | .55 |
| HR<sup>b</sup> | 86.0 (75.0, 97.0) | 87.0 (73.5, 95.5) | 85.5 (75.3, 98.0) | .65 |
| MAP (mm Hg)<sup>b</sup> | 93 (82.2, 103.0) | 91 (81.5, 104) | 94 (83, 103) | .66 |
| MAP ≤ 70 mm Hg | 13/260 (5.0%) | 6/89 (6.7%) | 7/171 (4.1%) | .35 |
| Hydroxychloroquine | 260 (92.9%) | 104 (97.2) | 156 (90.2) | .03 |
| Hydroxychloroquine plus azithromycin | 241 (86.1%) | 98 (91.6) | 143 (82.7) | .04 |
| Peripheral white cell count (X 10<sup>9</sup>/L)<sup>b</sup> | 7.3 (5.6, 10.2) (n=277) | 7.0 (5.7, 8.9) (n=106) | 7.6 (5.5, 11.1) (n=171) | .41 |
| Lymphocyte count (X 10<sup>9</sup>/L)<sup>b</sup> | 1.15 (0.78, 1.56) (n=260) | 1.14 (.84, 1.49) (n=102) | 1.20 (.77, 1.67) (n=158) | .62 |

<sup>a</sup> mean (± SD)
<sup>b</sup> median (interquartile range)
<sup>c</sup> Asian, Native American, Pacific Islander, or not identified

### Outcomes

Unadjusted outcomes for both groups are shown in Table 2. Overall mortality was significantly lower in the ivermectin group than in the usual care group (15.0% vs 25.2%, for ivermectin and usual care respectively, p=.03). Mortality was also lower for ivermectin treated patients in the subgroup of patients with severe disease (38.8% vs. 80.7%, p=.001). Differences in extubation rates between groups were not significant and there was also no difference in length of hospital stay.

**Table 2:** Univariate Clinical Outcomes by Treatment Group

|  | Number/total number (%) |  |  |  |  |
| --- | --- | --- | --- | --- | --- |
|  | Total (n=280) | Control (n=107) | Ivermectin (n=173) | OR (CI) | P value |
| **Mortality** |  |  |  |  |  |
| Total | 53 (18.9) | 27 (25.2) | 26 (15.0) | 0.52 (0.29 to 0.96) | .03 |
| Severe | 40/75 (53.3) | 21/26 (80.7) | 19/49 (38.8) | 0.15 (0.05 to 0.47) | .001 |
| Non-severe | 13/205 (6.3) | 6/81 (7.4) | 7/124 (5.6) | 0.75 (0.24 to 2.3) | .61 |
| Successful extubation | 17/62 (27.4) | 4/26 (15.4) | 13/36 (36.1) | 3.11 (0.88 to 11.00) | .07 |
| Length of stay (median, IQR) | 7.0 (4.0, 12.5) | 7.0 (4.0, 10.0) | 7.0 (4.0, 13.3) |  | .34 |

Abbreviations: IQR – interquartile range.

Univariate analysis found that patients dying with COVID-19 infection were older, had higher white blood cell counts and lower absolute lymphocyte counts (Table 3a) than survivors. Patients who died were also more likely to have been current or former smokers and have comorbid cardiac conditions, and were more likely to have severe pulmonary involvement and hypotension at study entry (Table 3b). In comparison to Caucasians, Blacks and Hispanics had lower odds of mortality in this cohort.

**Table 3a:** Univariate analysis of factors associated with mortality (continuous variables)

|  | Nonsurvivors | Survivors | Difference (CI) | P value |
| --- | --- | --- | --- | --- |
| Age, years<sup>a</sup> | 70.7 (15.1) | 57.0 (17.6) | 13.6 (8.5 to 18.8) | <.001 |
| Number of comorbidities<sup>a</sup> | 1.81 (1.49) | 1.63 (1.31) | 0.18 (-0.22 to 0.59) | .37 |
| BMI<sup>a</sup> | 28.3 (6.7) | 30.3 (8.0) | -2.0 (-4.4 to -0.33) | .09 |
| Peripheral white cell count (X 10<sup>9</sup>/L)<sup>b</sup> | 9.8 (6.1, 13.2) | 7.2 (5.5, 9.25) | 2.1 (0.8 to 3.6) | .001 |
| Lymphocyte count (X 10<sup>9</sup>/L)<sup>b</sup> | 0.77 (0.47, 1.15) | 1.29 (0.90, 1.68) | -0.46 (-0.63 to -0.30) | <.001 |

<sup>a</sup> mean ( ± SD)
<sup>b</sup> median (interquartile range)

**Table 3b:** Univariate analysis of factors associated with mortality (categorical variables)

|  | Mortality<br>Number/total<br>(percent) | OR (CI) | P value |
| --- | --- | --- | --- |
| Sex |  |  |  |
| Female | 24/127 (18.8) | 1.00 (0.55 to 1.83) | .99 |
| Male | 29/153 (19.0) | Reference |  |
| Smoking |  |  |  |
| Current or former | 18/46 (39.1) | 2.75 (0.14 to 0.55) | <.001 |
| Nonsmoker | 28/209 (13.9) | Reference |  |
| Race |  |  | .04 |
| White | 22/76 (28.9) | Reference |  |
| Black | 26/153 (17.0) | 0.50 (0.26 to 0.96) | .04 |
| Hispanic | 3/33 (9.1) | 0.24 (0.07 to 0.89) | .02 |
| Other<sup>a</sup> | 2/18 (11.1) | 0.31 (0.07 to 1.45) | .12 |
| Comorbidities |  |  |  |
| Diabetes | 19/90 (21.1) | 1.23 (0.66 to 2.30) | .52 |
| Cardiac | 15/43 (34.9) | 2.81 (1.37 to 5.75) | .004 |
| Pulmonary | 5/28 (17.9) | 0.92 (0.33 to 2.55) | .88 |
| Obesity | 20/114 (17.5) | 0.86 (0.46 to 1.59) | .62 |
| Renal | 6/24 (25.0) | 1.48 (0.56 to 3.94) | .43 |
| Cancer | 5/17 (29.4) | 1.87 (0.63 to 5.55) | .25 |
| Hypertension | 9/50 (18.0) | 0.93 (0.42 to 2.05) | .85 |
| Neurologic | 7/28 (25.0) | 1.49 (0.60 to 3.72) | .39 |
| Thyroid | 5/23 (21.7) | 1.21 (0.43 to 3.42) | .72 |
| Presentation<br>severity |  |  |  |
| Severe | 40/75 (53.3) | 16.88 (8.20 to 34.75) | <.001 |
| Non-severe | 13/192 (6.8) |  |  |
| MAP ≤ 70 | 8/13 (38.5) | 9.08 (2.82, 29.28) | <.001 |
| Hydroxychloroquine | 50/260 (19.2) | 1.35 (0.38 to 4.78) | .64 |
| Hydroxychloroquine<br>plus azithromycin | 47/241 (19.5) | 1.33 (0.53 to 3.37) | .54 |

<sup>a</sup>Asian, Native American, Pacific Islander, or not identified

In the multivariate analysis, adjusting for demographic factors, between-group differences in mortality risks, and concomitant use of hydroxychloroquine (with or without azithromycin), independent predictors of in-hospital mortality included treatment group, age, severe pulmonary disease category, and reduced lymphocyte count (Table 4). Similarly, the Cox regression showed ivermectin was associated with a significantly lower hazard ratio for mortality of 0.37 (CI 0.19 - 0.70, p=.003). Complete case analysis on the entire cohort without missing data was similar (HR 0.40, 0.22- 0.74), p=.003).

**Table 4:** Multivariate analysis of factors associated with mortality

|  | OR (CI) | P value |
| --- | --- | --- |
| Treatment group: |  |  |
| Ivermectin | 0.27 (0.09, 0.85) | .03 |
| Control | Reference |  |
| Age | 1.05 (1.02, 1.09) | .005 |
| Female sex | 0.69 (0.25, 1.91) | .48 |
| Current or former smoker | 3.43 (0.94, 12.48) | .06 |
| Race |  | .165 |
| Black | 0.66 (0.22, 1.99) | .464 |
| Hispanic | 0.14 (0.02, 1.10) | .061 |
| Other | 0.77 (0.06, 9.88) | .843 |
| Comorbidities |  |  |
| Diabetes | 1.13 (0.38, 3.36) | .83 |
| Cardiac | 1.62 (0.45, 5.83) | .46 |
| Pulmonary | 0.15 (0.17, 1.24) | .08 |
| Hypertension | 1.00 (0.22 to 4.56) | .99 |
| BMI | 0.99 (0.90, 1.08) | .77 |
| Severe presentation | 26.88 (8.11, 89.05) | <.001 |
| MAP ≤ 70 mm Hg | 2.10 (0.26, 7.04) | .73 |
| Peripheral white cell count | 1.09 (.958, 1.24) | .19 |
| Lymphocyte count | 4.04 (1.35, 12.09) | .01 |

## Discussion

In this multihospital retrospective cohort study, we observed a significant association with ivermectin on survival for patients admitted with Covid-19. This association was also seen in the subset of patients with severe pulmonary disease.

Similar to other studies, we noted that older age, cardiac disease, current or former smoking, more severe pulmonary involvement at presentation, higher white blood cell counts, and lower lymphocyte counts emerged as risk markers for in-hospital mortality. The overall mortality, and mortality in intubated patients, in our usual care group was similar to what was reported in previous studies. Richardson et al reported an overall mortality of 21% in their New York City cohort, with a mortality of 88% in intubated patients.^5^ Fei Zhou et al reported a 28.2% mortality in their cohort of hospitalized patients in Wuhan, China; their intubated patients had a mortality of 96.9%.^6^ In contrast to Mehra et al, we did not see a mortality effect for hydroxychloroquine, with or without the addition of azithromycin.^7^ This may have been due to the small number of patients who were not treated with these agents; our study was underpowered to detect a difference. We also hypothesize that precautionary measures in the hospitals’ protocol for hydroxychloroquine use could have prevented them from developing fatal arrhythmias. These included baseline EKG and daily QT monitoring by telemetry for any patient receiving hydroxychloroquine or combination therapy, avoidance of azithromycin if patient’s baseline QTc was greater than 460msec, and discontinuation of hydroxychloroquine if there was a concerning elevation in QTc or if the patient’s cardiologist recommended discontinuation.

We did not confirm an increase risk of mortality in Blacks. This was likely due to difference in age; white patients were significantly older (66.8 vs 59.1 years; mean difference 7.7 years, CI 3.0 - 12.4, p=.001).

In the Ivermectin group, thirteen patients received a second dose of Ivermectin (200 mcg/kg) on day 7, since they were still hospitalized. Due to the low numbers, we did not perform any further analysis of the redosing. We also did not observe any significant side-effect from Ivermectin use.

We did not observe a significant difference in hospital length of stay between the groups (median 7 days for both groups). Possible explanation could include delay in discharging patients to other facilities (skilled nursing facilities, inpatient rehabs, etc) due to lag in obtaining required repeat COVID testing results.

Need for mechanical ventilation was not adopted as outcome of interest, as national guidelines and practice patterns for intubation criteria changed throughout the length of the study.

We did not find a lower mortality in the non-severe patients treated with ivermectin; however, our study was not powered to assess these differences as the overall mortality in non-severe patients was low. Similarly, the study was not powered to determine whether extubation rates were higher in the Ivermectin group. These should be investigated further with a larger randomized controlled trial.

Our study has several limitations. Due to the retrospective observational nature of the study, despite adjustment for known cofounders, we cannot exclude the possibility of unmeasured confounding factors. Although more of the control group was enrolled earlier in the study, suggesting the possibility of timing bias, this may be offset by preferential treatment of more severe patients with ivermectin early in the study due to low initial availability. We also did not find consistently better mortality outcomes with time over the short duration of this study. Most of our patients studied received hydroxychloroquine with or without azithromycin and we are unable to determine whether these medications had an added benefit, or whether mortality would have been better in both groups without these agents.

We have shown that ivermectin administration was significantly associated with lower mortality among patients with COVID-19, particularly in patients with more severe disease. Interpretation of these findings are tempered by the limitations of the retrospective design and the possibility of confounding. Appropriate dosing for this indication is not known; nor are the effects of ivermectin on viral load, or in patients with milder disease. Further studies in appropriately designed randomized trials are recommended before any conclusions can be made.

## Data Availability

De-dentified data is available upon request and approval by the Broward Health IRB.

## Acknowledgements

Dr. Edward Gracely, Ph.D. for his support with the statistical analysis.

